# Low adherence to existing model reporting guidelines by commonly used clinical prediction models

**DOI:** 10.1101/2021.07.21.21260282

**Authors:** Jonathan H. Lu, Alison Callahan, Birju S. Patel, Keith E. Morse, Dev Dash, Nigam H. Shah

**Affiliations:** Center for Biomedical Informatics Research, Stanford University School of Medicine, Stanford, CA; Department of Pediatrics, Stanford University School of Medicine, Stanford, CA, USA; Department of Clinical Informatics, Lucile Packard Children’s Hospital, Palo Alto, CA, USA

**Keywords:** electronic health records, clinical trials, Research Report / standards, machine learning, Decision Support Techniques

## Abstract

**Objective:** To assess whether the documentation available for commonly used machine learning models developed by an electronic health record (EHR) vendor provides information requested by model reporting guidelines.

**Materials and Methods:** We identified items requested for reporting from model reporting guidelines published in computer science, biomedical informatics, and clinical journals, and merged similar items into representative “atoms”. Four independent reviewers and one adjudicator assessed the degree to which model documentation for 12 models developed by Epic Systems reported the details requested in each atom. We present summary statistics of consensus, interrater agreement, and reporting rates of all atoms for the 12 models.

**Results:** We identified 220 unique atoms across 15 model reporting guidelines. After examining the documentation for the 12 most commonly used Epic models, the independent reviewers had an interrater agreement of 76%. After adjudication, the model documentations’ median completion rate of applicable atoms was 39% (range: 31%-47%). Most of the commonly requested atoms had reporting rates of 90% or above, including atoms concerning outcome definition, preprocessing, AUROC, internal validation and intended clinical use. For individual reporting guidelines, the median adherence rate for an entire guideline was 54% (range: 15%-71%). Atoms reported half the time or less included those relating to fairness (summary statistics and subgroup analyses, including for age, race/ethnicity, or sex), usefulness (net benefit, prediction time, warnings on out-of-scope use and when to stop use), and transparency (model coefficients).

Atoms relating to reliability also had low reporting, including those related to missingness (missing data statistics, missingness strategy), validation (calibration plot, external validation), and monitoring (how models are updated/tuned, prediction monitoring).

**Conclusion:** There are many recommendations about what should be reported about predictive models used to guide care. Existing model documentation examined in this study provides less than half of applicable atoms, and entire reporting guidelines have low adherence rates. Half or less of the reviewed documentation reported information related to usefulness, reliability, transparency and fairness of models. There is a need for better operationalization of reporting recommendations for predictive models in healthcare.

**KEY POINTS:** *Question:* How often does documentation for commonly deployed clinical predictive models report the information requested by model reporting guidelines?

*Finding:* Combining the recommendations from 15 model reporting guidelines, we identified 220 unique requested items. We reviewed the documentation of 12 commonly deployed Epic models and assessed the completion rate of applicable items. The median completion rate was 39%. While the most commonly requested items were highly reported, information on usefulness, reliability, transparency and fairness was missing from at least half of documentation.

*Meaning:* There is incomplete documentation for model users to ensure that deployed models are useful, reliable, transparent and fair.

## INTRODUCTION

Despite good predictive performance in metrics such as the area under the receiver operating characteristic (AUROC) curve, the use of machine learning models trained on electronic health records (EHR) data^1^ to guide care does not always translate into clinical gains in the form of better medical care, lower cost or more equitable outcomes,^2–4^ leading to a gap referred to as an “Artificial Intelligence (AI) chasm”.^5^ Some potential causes of this chasm are that current models are not useful,^4,6,7^ reliable,^8,9^ or fair.^10–18^ Nevertheless, predictive models have been deployed in healthcare settings without transparency or independent validation,^19,20^ and their subsequent failures have been met with public outcry.^2,21–23^

Adhering to model reporting guidelines is one way to improve the usefulness,^24–28^ fairness,^29,30^ and reliability^27,31–34^ of clinical predictive models. Reporting guidelines have long been used to assess the strength of clinical trial studies,^35,36^ observational studies,^37^ and diagnostic studies.^38^ Guidelines concerning predictive models are receiving increasing attention, including from the National Institutes of Health,^39^ and several more are in development.^40–42^

While there has been increasing interest in model reporting guidelines, the degree to which currently deployed models adhere to these guidelines has not been studied. One review examining 164 models described in the scientific literature found low reporting rates of demographic variables such as race (36%) and socioeconomic status (8%) as well as low external validation rates (12%).^43^ A critical review of published models for diagnosis and prognosis of COVID-19 found that most models were at high risk of bias due to poor reporting.^44^

The purpose of this analysis is to assess whether the documentation available for commonly deployed models provides the information requested by model reporting guidelines. Compared to previous work,^43,44^ we focus on user-facing product documentation accompanying models. Thus, we are able to analyze models that have been deployed in practice but not yet described in peer-reviewed publications. Furthermore, we make a comprehensive assessment of the reporting rates of every requested item in the guidelines.

## METHODS

We searched MEDLINE via PubMed using queries for “machine learning model card” and “reporting machine learning” in November 2020. We reviewed citations to find additional publications. Finally, we excluded publications that did not give specific model reporting recommendations. We included all Explanation and Elaboration documents, AI-specific extensions and multi-part guidelines for papers which had them.

We gathered the set of reportable items in these reporting guidelines and deduplicated these items; i.e. we merged similar items into distinct, representative “atoms.” For example, “report the intended user of the model”^24^ or “describe external validation strategy”^31^ are unique atoms. We performed the de-duplication in two rounds. First, we created an initial set of atoms by reviewing each reporting guideline, including the Explanation & Elaboration documents and AI-extensions to verify that every publication’s atoms were captured. Second, we reviewed each atom and merged those that requested the same information. We recorded the phrases describing the atoms to enable a full traceback of which items were merged to the same atom. Lastly, we created a one-line summary of each atom to share in our reported results. To facilitate summarization, we mapped the atoms (eFile 1) to several general stages in the creation and evaluation of a machine learning model to guide care (Figure 4 of Jung et al^7^, eFigure 1). The general stages are Use Case, Model Formulation, Model Development, Fairness in Model Development, Practical Feasibility, Utility Assessment, Deployment Design, Deployed Model (including Execution and Workflow) and Prospective Evaluation. Each stage has a color (eFigure 1), so our tables and figures use the stage’s color for atoms in that stage. We also mapped atoms to specific tasks in the model development process (eFile 1).

We next identified the model documentation to review. Epic provides Cognitive Computing Model Briefs (hereafter referred to as Model Briefs), which are user-facing documentation sheets (analogous to a drug package insert) for models available from the vendor. Each Model Brief has a community adoption score which represents the number of organizations that have used a specific model as a proportion of the number of organizations using any model, and takes values from a scale ranging from 1 to 3. We chose all models that had a community adoption score of 2 or 3 in March 2021. The six Model Briefs with community adoption score 3 out of 3, downloaded on March 8th, 2021, were for Deterioration Index, Early Detection of Sepsis, Risk of Unplanned Readmission (Version 2), Risk of Patient No-Show (Version 2), Pediatric Risk of Hospital Admission or ED Visit (Version 2), and Risk of Hospital Admission or ED Visit. The six Model Briefs with community adoption 2 out of 3, downloaded on April 13th, 2021, were for Inpatient Risk of Falls, Projected Block Utilization, Remaining Length of Stay, Risk of Hospital Admission for Heart Failure, Risk of Hospital Admission or ED Visit for Asthma, and Risk of Hypertension.

Four reviewers read each of the 12 Model Briefs and assessed whether they reported information specified in the atoms (eMethods). Specifically, for each atom, each reviewer first determined if the atom was applicable to the model. For example, an atom such as “A link to the clinical trial registration” is not applicable to models where documentation does not intend to describe a clinical trial. When atoms were applicable, the reviewer decided whether the Model Brief reported the information requested in the atom.

Atoms had consensus when all four reviewers agreed that an atom was reported by the Model Brief, was not reported by the Model Brief, or was determined to be not applicable. For atoms that did not have consensus across all four reviewers, a designated adjudicator reviewed the atoms and the corresponding Model Brief content, to independently adjudicate the reviewer responses.

To determine the inter-rater agreement, we calculated the fraction of atoms that a pair of reviewers agreed were reported, were not reported, or were determined to be not applicable, averaged across all Model Briefs and pairs of the four reviewers.

To standardize nomenclature, we define that an atom is “requested” by a reporting guideline if any reportable item from the reporting guideline was merged into that atom. We define that an atom is “reported” by a Model Brief if we determine that the Model Brief contained the information requested in the atom, after adjudication.

An atom’s reporting rate is the number of Model Briefs that reported the atom divided by the number of Model Briefs for which the atom was applicable. A Model Brief’s completion rate of a given group of atoms is the number of atoms reported by the Model Brief divided by the number of atoms that were applicable to that Model Brief. Finally, the adherence rate to a reporting guideline is the completion rate of atoms requested by the specific reporting guideline, averaged across all Model Briefs. We calculate median, interquartile range (IQR) and range for atoms’ reporting rates, Model Briefs’ completion rates, and reporting guidelines’ adherence rates, as appropriate.

## RESULTS

### Atoms Requested by Model Reporting Guidelines

Our MEDLINE search resulted in an initial list of 26 publications.^24–30,38,41,45–62^ We reviewed citations and found 3 additional publications.^32–34^ We excluded publications that did not give specific model reporting recommendations to arrive at our final list of 15 model reporting guidelines published (Table 1).^24–36,38,45–47,62–65^

**Table 1:** Summary of 15 Model reporting guideline papers. “Total citations” sums the citations for each of the papers, excluding the Explanation and Elaboration papers. “Atoms” indicates the number of deduplicated atoms sourced from that guideline. We included the Explanation and Elaboration papers for CONSORT, SPIRIT, TRIPOD and PROBAST [^32,63–65^]. For CONSORT and SPIRIT, we also included the AI-specific extensions ^25,26^. We grouped Risk Prediction Models II ^31^ with the Risk Prediction Models I paper ^62^.

| Abbreviation | Title | Author and Year | Journal | Total citations* | Atoms |
| --- | --- | --- | --- | --- | --- |
| CONSORT-AI | CONSORT 2010 statement: updated guidelines for reporting parallel group randomised trials<br>CONSORT 2010 Explanation and Elaboration: updated guidelines for reporting parallel group randomised trials<br>Reporting guidelines for clinical trial reports for interventions involving artificial intelligence: the CONSORT-AI extension | Schulz 2010<br>Moher 2010<br>Liu 2020 | Lancet<br>Journal of Clinical Epidemiology<br>Nature | 11529 | 68 |
| Risk | Risk prediction models: I. Development, internal validation, and assessing the incremental value of a new (bio)marker<br>Risk prediction models: II. External validation, model updating, and impact assessment | Moons 2012<br>Moons 2012 | Heart<br>Heart | 1320 | 41 |
| SPIRIT-AI | SPIRIT 2013 Statement: Defining Standard Protocol Items for Clinical Trials<br>SPIRIT 2013 explanation and elaboration: guidance for protocols of clinical trials<br>Guidelines for clinical trial protocols for interventions involving artificial intelligence: the SPIRIT-AI extension | Chan 2013<br>Chan 2013<br>Rivera 2020 | Annals of Internal Medicine<br>BMJ<br>Nature | 2952 | 75 |
| ABCD | Towards better clinical prediction models: seven steps for development and an ABCD for validation | Steyerberg 2014 | European Heart Journal | 709 | 33 |
| CHARMS | Critical Appraisal and Data Extraction for Systematic Reviews of Prediction Modelling Studies: The CHARMS Checklist | Moons 2014 | PLOS Medicine | 565 | 63 |
| TRIPOD | Transparent Reporting of a multivariable prediction model for Individual Prognosis Or Diagnosis (TRIPOD): The TRIPOD Statement<br>Transparent Reporting of a multivariable prediction model for Individual Prognosis Or Diagnosis (TRIPOD): Explanation and Elaboration | Collins 2015<br>Moons 2015 | Annals of Internal Medicine,<br>Annals of Internal medicine | 3031 | 86 |
| STARD | STARD 2015 guidelines for reporting diagnostic accuracy studies: explanation and elaboration | Cohen 2016 | BMJ Open | 711 | 55 |
| Guidelines | Guidelines for Developing and Reporting Machine Learning Predictive Models in Biomedical Research: A Multidisciplinary View | Luo 2016 | Journal of Medical Internet Research | 244 | 49 |
| ML Test Score | The ML Test Score: A Rubric for ML Production Readiness and Technical Debt Reduction | Breck 2017 | IEEE International Conference on Big Data | 68 | 34 |
| PROBAST | PROBAST: A Tool to Assess the Risk of Bias and Applicability of Prediction Model Studies<br>PROBAST: A Tool to Assess Risk of Bias and Applicability of Prediction Model Studies: Explanation and Elaboration | Moons 2019<br>Moons 2019 | Annals of Internal Medicine<br>Annals of Internal medicine | 284 | 55 |
| Model Cards | Model Cards for Model Reporting | Mitchell 2019 | ACM Fairness, Accountability and Transparency | 311 | 49 |
| Model Facts Labels | Presenting machine learning model information to clinical end users with model facts labels | Sendak 2020 | npj Digital Medicine | 14 | 37 |
| MINIMAR | MINIMAR (MINimum Information for Medical AI Reporting): developing reporting standards for artificial intelligence in health care | Hernandez-Boussart 2020 | JAMIA | 18 | 28 |
| MI-CLAIM checklist | Minimum information about clinical artificial intelligence modeling: the MI-CLAIM checklist | Norgeot 2020 | Nature Medicine | 24 | 40 |
| Trust and Value Checklist | AI-Enabled Clinical Decision Support Software: A “Trust and Value Checklist” for Clinicians | Silcox 2020 | NEJM Catalyst | 2 | 26 |

Publication venues include computer science venues (ACM Fairness, Accountability, and Transparency^29^ and IEEE^27^), biomedical informatics journals (Journal of the American Medical Informatics Association, npj Digital Medicine, Journal of Medical Internet Research^24,30,45^), and clinically-focused journals (Annals of Internal Medicine, BMJ, Nature Medicine, Heart, European Heart Journal, PLOS Medicine, and NEJM Catalyst^26,28,31–35,38,46,47,62^). Four guidelines published between 2010 and 2015 have been cited by other articles over 1000 times to date, while four guidelines were published after 2019 and have been cited less than 50 times to date.

Of the 15 reporting guidelines, 11 had examples of how to complete their requested atoms.^24,27,29,30,32,33,38,62–65^ However, only 5 showed a full example completing all atoms for a single model,^24,29,30,33,62^ and only 1 of those models was deployed in a health system.^24,66^

After deduplication, there were 220 distinct atoms requested by all of the reporting guidelines (eFile 1). We provide a cross tabulation of the 220 atoms against the 15 reporting guidelines (eTable 1) to show the most relevant guideline for a task. For example, the TRIPOD reporting guideline has more atoms requesting details on preprocessing^47^ while MI-CLAIM has more atoms requesting details for model examinations.^46^

Figure 1 summarizes the model reporting guidelines in terms of the number of atoms that map to each stage in the creation and evaluation of a machine learning model (Figure 4 of Jung et al^7^, eFigure 1). This allows selecting the stage-appropriate reporting guideline: for example, Model Cards^29^ contributes the most atoms to fairness in model development, while Model Facts Labels^24^ or CONSORT-AI^25^ contribute the most atoms to use case assessment.

**Figure 1:**
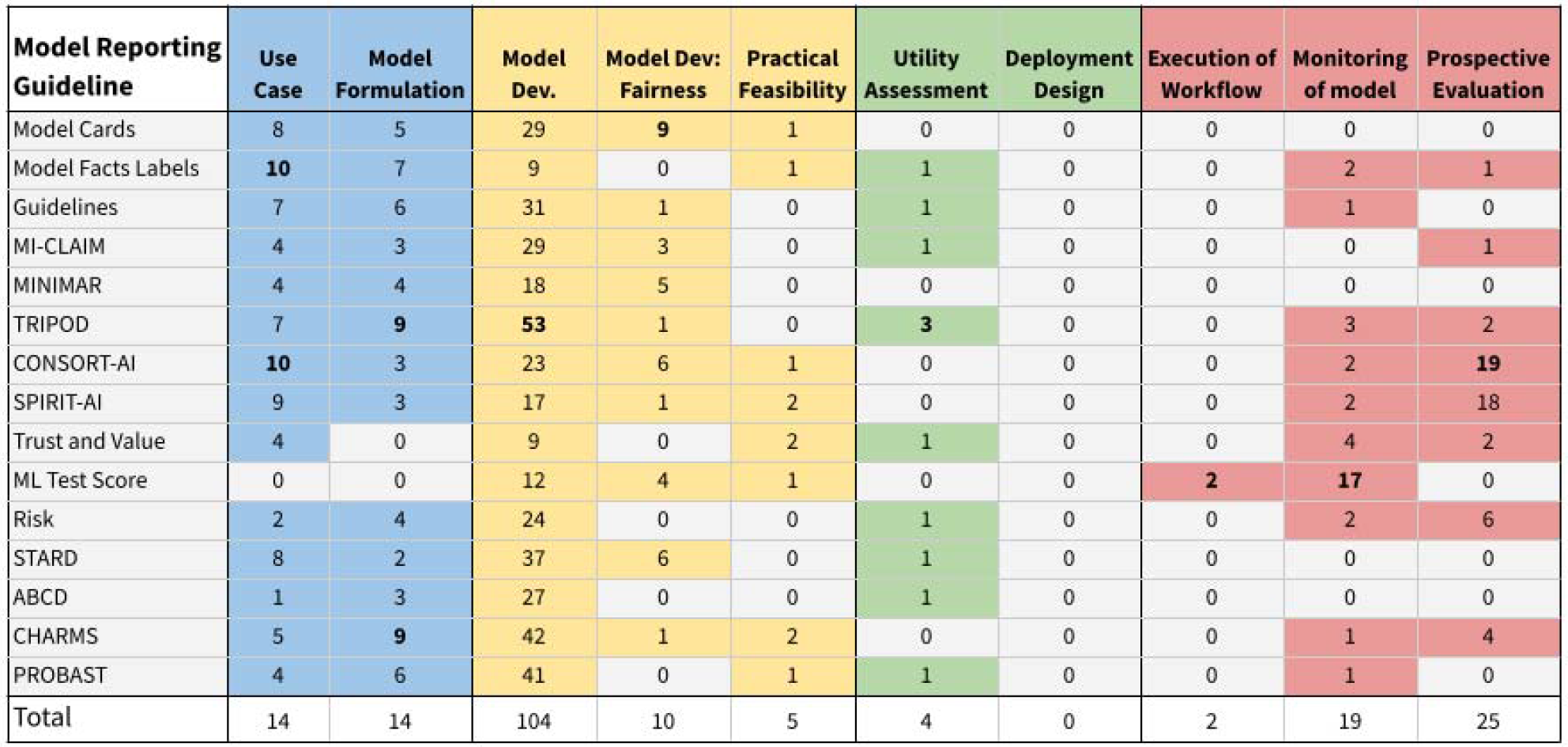
Model reporting guidelines (in rows), with their atoms mapped onto different stages in the creation and evaluation of a machine learning model to guide care. ^7^ Stages are listed in eFigure 1. Each cell is the number of atoms contributed by the relevant model reporting guideline towards a given stage of the workflow (columns). Model Dev. stands for Model Development. The highest number in each column is bolded.

There are stages in the creation and evaluation of a machine learning model for which reporting guidelines focus less; for example, there are less than five atoms related to Deployment Design, e.g.. considering work capacity and resources to perform interventions, and for Utility Assessment, e.g. considering the net benefit of taking actions guided by the model’s output. Meanwhile, the Model Development step comprises 53 atoms.

Table 2 shows the atoms requested by at least 10 out of the 15 reporting guidelines. The most commonly requested atoms relate to model development tasks, such as preprocessing, missing data handling, model performance including handling of uncertainty (e.g. confidence intervals, statistical significance) or AUROC, and internal validation. A total of 28 distinct performance metrics were requested (eTable 2), including discrimination, calibration, classification, goodness-of-fit, utility, and comparisons of model discrimination.

**Table 2:**
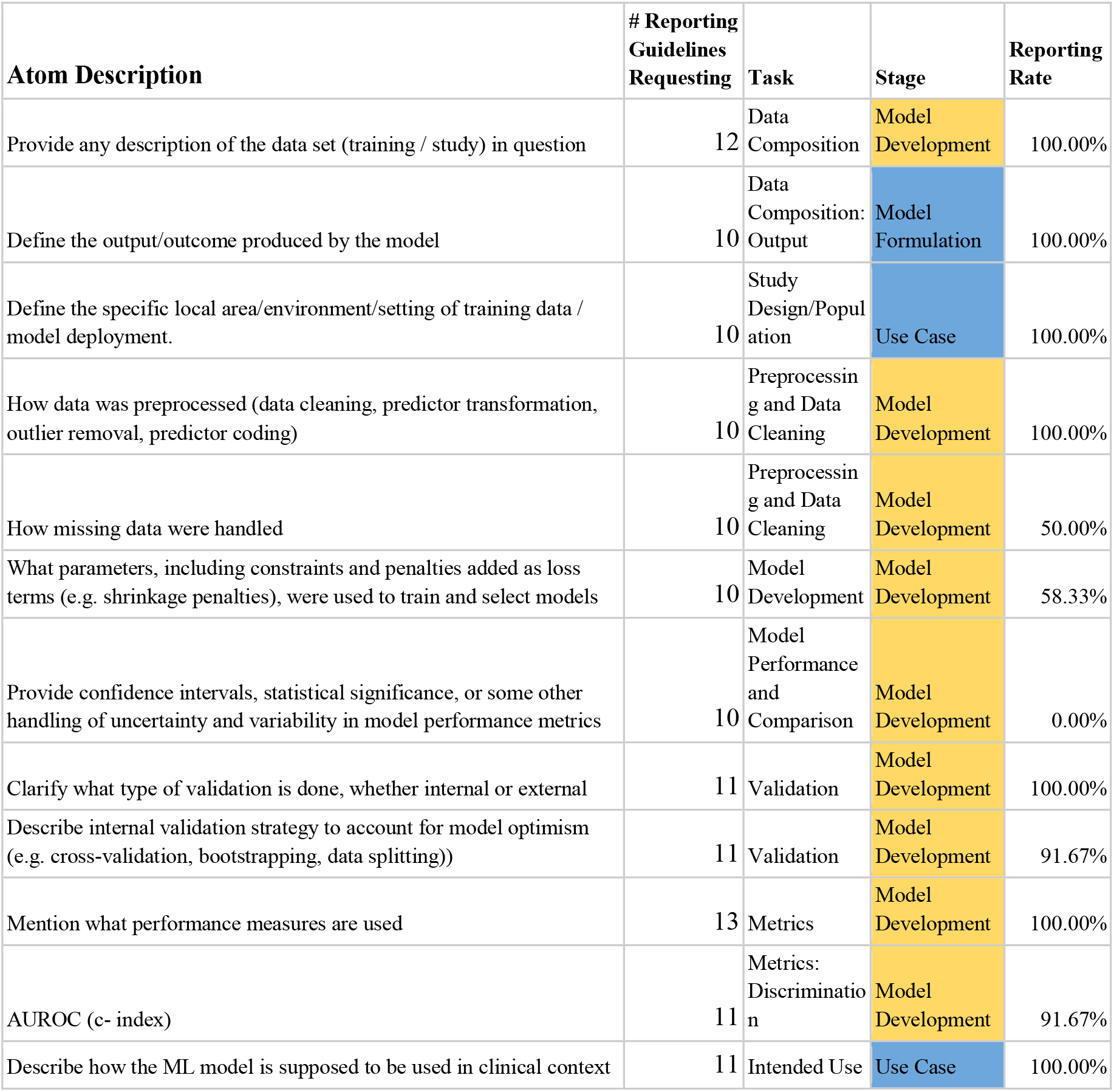
Commonly Requested Atoms across reporting guidelines. This table lists all atoms requested by at least 10 model reporting guidelines. Reporting Rate indicates the % of the Model Briefs that reported the information requested in the atom. Task and Stage indicate the atoms’ related task and related stage of clinical predictive model development, respectively ^7^.

Finally, there were 77 atoms that were requested by just one reporting guideline (eTable 3). ML Test Score had 20 unique atoms related to model deployment and monitoring, such as steps for model updating and rollback. CONSORT-AI and SPIRIT-AI had a combined 21 clinical trial-specific atoms, which mostly did not apply to Epic’s Model Briefs (e.g. random allocation methods). Twelve uniquely requested atoms were model performance metrics such as the F-Score or Relative Utility.

### Reporting of deduplicated atoms by Model Briefs

A median of 93 (IQR: 88-95, range: 66-108) atoms per brief underwent adjudication for discordant findings by reviewers. Interrater agreement on atom reporting was 76%.

There were 40 commonly reported atoms, whose information was reported by over 90% of the Model Briefs (eTable 4). These atoms requested information about model development and formulation, including the training data set, preprocessing, model type, internal validation, and performance metrics. These 40 commonly reported atoms by Model Briefs included 9 of the 12 most commonly requested atoms across the reporting guidelines (Table 2).

There were 75 rarely reported atoms, whose information was reported in less than 10% of the Model Briefs (eTable 5). These atoms included missing data statistics, blinding of predictor/outcome assessors, variability of performance measures (e.g. confidence intervals), reporting of model coefficients or most predictive features, model examinations including performance errors and intersectional subgroup analyses, user-facing materials and warnings on when to stop use of model, and monitoring of input data and model predictions. In addition, of 28 distinct performance metrics requested, only AUROC (92%), PPV (67%), and sensitivity (42%) were reported by more than a fifth of the Model Briefs (eTable 2).

There were 34 atoms for which reviewers had no consensus across any of the 12 Model Briefs (eTable 6). These atoms without consensus included atoms related to data collection, reference standards, and performance metrics, where there was disagreement about applicability. Of 220 atoms, 176 (80%) were considered applicable for at least one Model Brief. Of these 176, 119 (68%) were reported by at least one Model Brief. Atoms had a median reporting rate across briefs of 25% (IQR: 0%-83%, range: 0%-100%).

From the standpoint of a specific Model Brief (eTable 7), there were a median of 171 applicable atoms (IQR: 170-173, range: 166-173) per brief of which a median of 67 atoms were reported in the brief (IQR: 64-74, range: 53-81). A Model Brief’s median completion rate of applicable atoms was 39% (IQR: 37%-43%, range: 31%-47%). After excluding all atoms corresponding to performance metrics -- to ensure briefs were not penalized for not reporting multiple redundant performance metrics -- the median completion rate for applicable atoms was 43% (IQR: 41%-48%, range: 33%-52%). Lastly, every Model Brief covered the following use case-related atoms: how the model is to be used in clinical care, who will use the model, ways the model could impact clinical care, and rationale for use.

### Adherence to Entire Reporting Guidelines by Model Briefs

Table 3 shows the adherence rates to individual reporting guidelines, which is the Model Briefs’ average completion rate of atoms requested by the reporting guideline. Model reporting guidelines had a median adherence rate of 53% (IQR: 50%-63%, range: 18%-74%). The ML Test Score had the lowest adherence rate (18%) while Model Facts Labels had the highest (74%). After excluding redundant performance metrics as before, the median adherence rates remained similar, at 57% (IQR: 50%-70%, range: 16%-73%).

**Table 3:** Adherence rates to entire reporting guidelines across Model Briefs. Cells are colored green if above 50%, yellow if between 25% and 50%, and red if below 25%. The AVERAGE, MIN, and MAX columns are the average, minimum and maximum adherence rates for the model reporting guidelines, respectively.

|  | EPIC MODEL BRIEFS |  |  |  |  |  |  |  |  |  |  |  |  |  |  |  |  |
| --- | --- | --- | --- | --- | --- | --- | --- | --- | --- | --- | --- | --- | --- | --- | --- | --- | --- |
| MODEL REPORTING GUIDELINES | Deterioration Index | Early Detection of Sepsis | Risk of Unplanned Readmission | Risk of Patient No-Show | Pediatric Risk of Hospital Admission or ED Visit | Risk of Hospital Admission or ED Visit | Inpatient Risk of Falls | Projected Block Utilization | Remaining Length of Stay | Risk of Admission of Heart Failure | Risk of Hospital Admission or ED Visit for Asthma | Risk of Hypertension | AVERAGE | MIN | MAX | Applicable Atoms (Average) | Applicable Atoms (Range) |
| Model Cards | 66% | 47% | 63% | 51% | 40% | 69% | 51% | 45% | 47% | 47% | 41% | 57% | 52% | 40% | 69% | 48.7 | [47, 49] |
| Model Facts Labels | 77% | 71% | 80% | 89% | 71% | 80% | 71% | 71% | 77% | 60% | 63% | 71% | 74% | 60% | 89% | 34.9 | [34, 35] |
| Guidelines | 64% | 66% | 66% | 66% | 57% | 74% | 62% | 49% | 66% | 64% | 64% | 66% | 64% | 49% | 74% | 47.0 | [47, 47] |
| MI-CLAIM | 55% | 58% | 63% | 58% | 47% | 68% | 53% | 34% | 47% | 53% | 45% | 58% | 53% | 34% | 68% | 38.0 | [38, 38] |
| MINIMAR | 71% | 71% | 79% | 61% | 68% | 86% | 71% | 46% | 61% | 75% | 61% | 82% | 69% | 46% | 86% | 28.0 | [28, 28] |
| TRIPOD | 63% | 63% | 61% | 48% | 42% | 61% | 47% | 36% | 55% | 48% | 44% | 51% | 51% | 36% | 63% | 75.5 | [72, 77] |
| CONSORT-AI | 63% | 43% | 63% | 60% | 33% | 67% | 53% | 47% | 47% | 49% | 42% | 51% | 52% | 33% | 67% | 42.4 | [40, 43] |
| SPIRIT-AI | 61% | 55% | 54% | 54% | 38% | 61% | 44% | 49% | 51% | 41% | 39% | 46% | 49% | 38% | 61% | 40.4 | [38, 41] |
| Trust and Value | 46% | 33% | 39% | 50% | 29% | 42% | 38% | 46% | 46% | 25% | 33% | 46% | 39% | 25% | 50% | 23.9 | [23, 24] |
| ML Test Score | 27% | 15% | 33% | 24% | 9% | 33% | 15% | 6% | 18% | 12% | 9% | 15% | 18% | 6% | 33% | 32.9 | [32, 33] |
| Risk | 64% | 65% | 63% | 53% | 50% | 68% | 53% | 48% | 56% | 56% | 56% | 56% | 57% | 48% | 68% | 33.7 | [32, 34] |
| STARD | 54% | 45% | 50% | 40% | 29% | 52% | 52% | 39% | 34% | 40% | 40% | 52% | 44% | 29% | 54% | 48.8 | [46, 50] |
| ABCD | 65% | 65% | 48% | 55% | 61% | 68% | 52% | 39% | 55% | 65% | 61% | 61% | 58% | 39% | 68% | 31.0 | [31, 31] |
| CHARMS | 78% | 70% | 68% | 65% | 56% | 75% | 66% | 47% | 70% | 65% | 63% | 64% | 66% | 47% | 78% | 55.0 | [53, 56] |
| PROBAST | 69% | 71% | 67% | 62% | 53% | 68% | 58% | 46% | 60% | 60% | 58% | 60% | 61% | 46% | 71% | 52.2 | [49, 53] |

### Requested, but Less Reported Atoms

We identified 29 atoms that were requested by at least 4 out of 15 the reporting guidelines, but were reported by 50% or less of Model Briefs (Table 4). Many of these less reported atoms are related to fairness, i.e. data set representativeness and performance across subgroups. These include summary statistics of key characteristics of the training data set (reporting rate 50%) or disaggregating performance by a subgroup (33%). Key factors such as age (50%), sex (33%), and other relevant factors (50%) lacked both summary statistics and disaggregated performance.

**Table 4:**
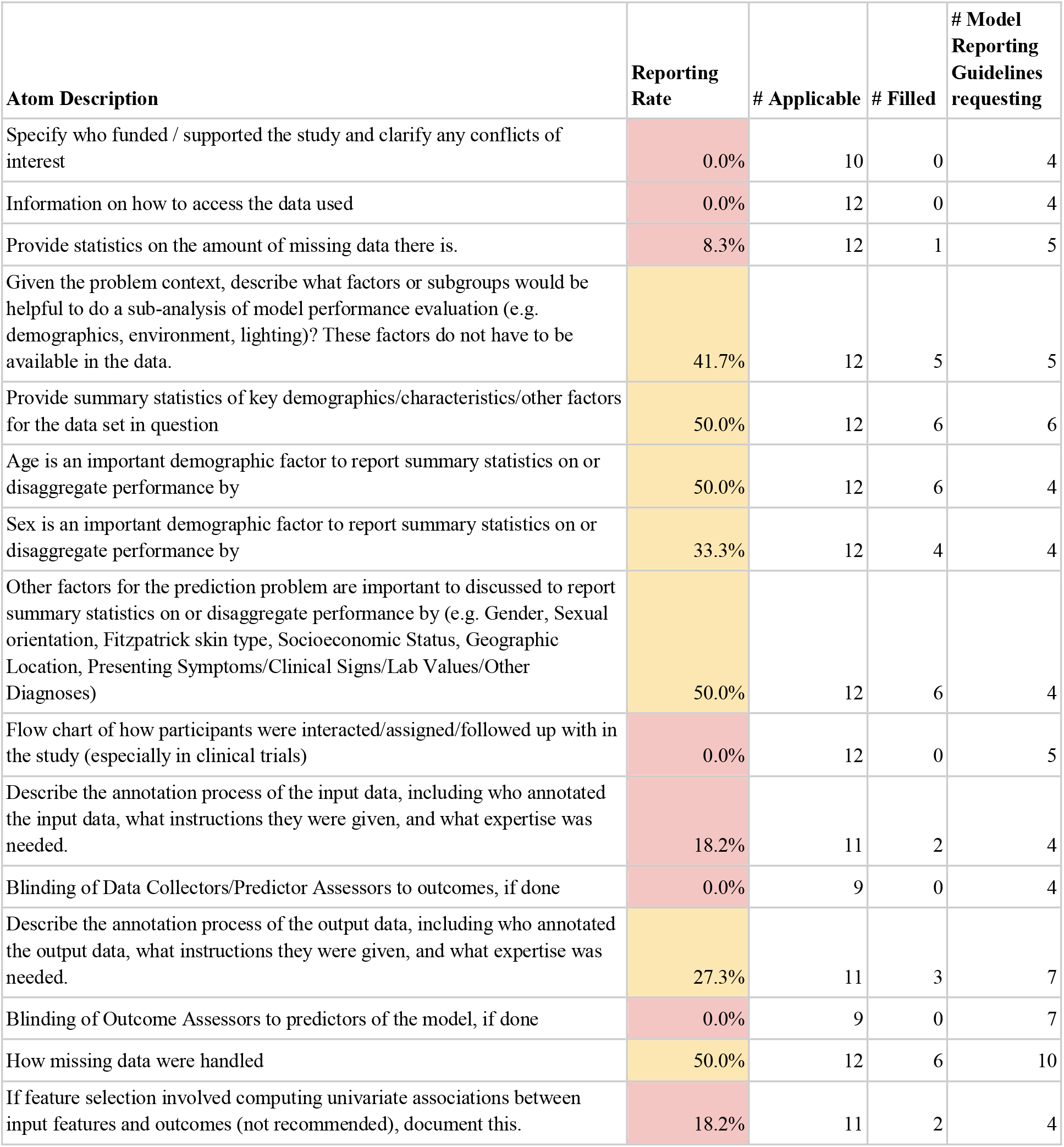
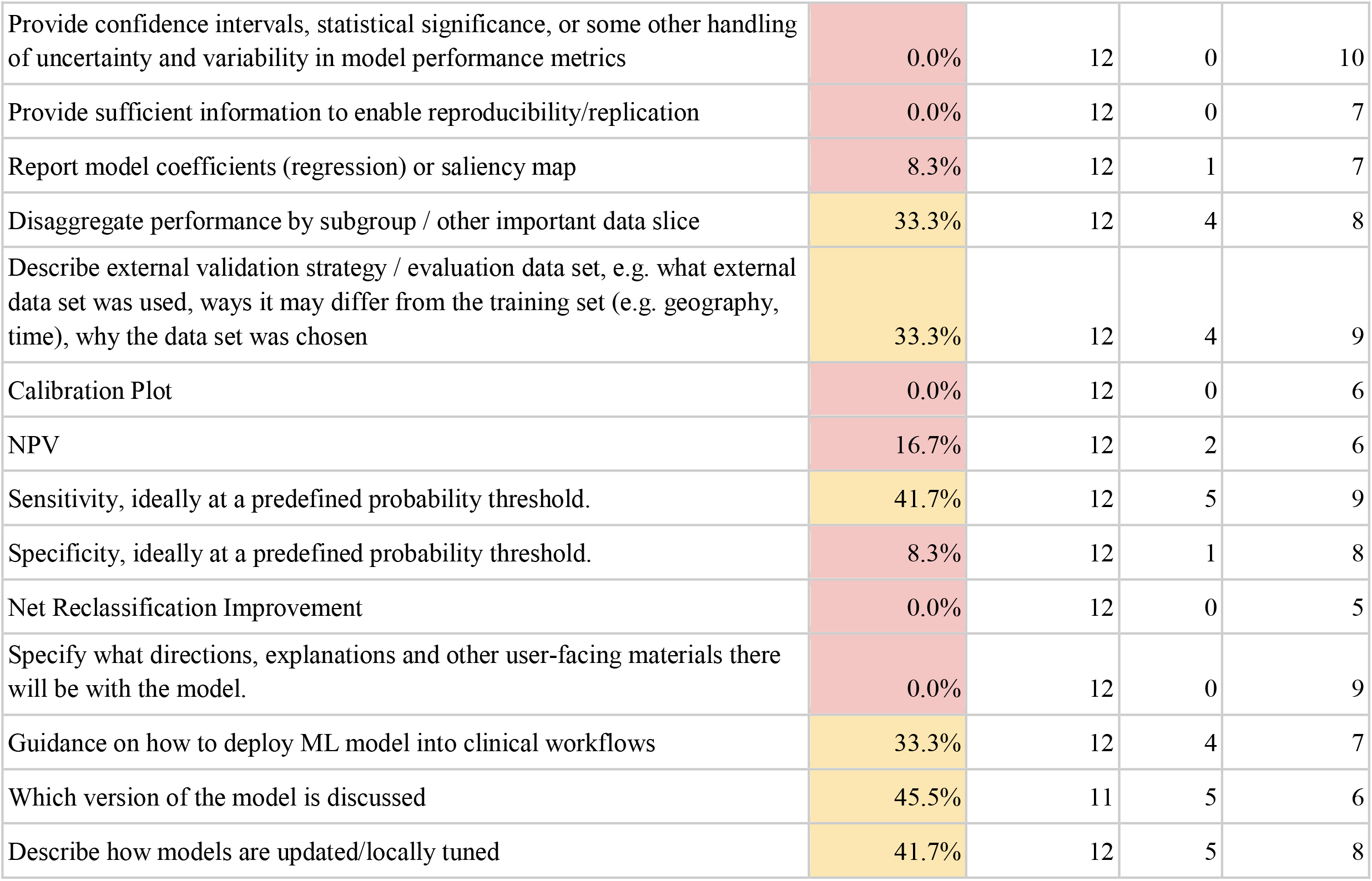
Requested, but less Reported atoms. All atoms requested by 4 or more model reporting guidelines but reported by no more than 50% of applicable Model Briefs are listed. The “Reporting Rate” column is colored yellow if between 25% and 50%, and red if below 25%.

| **Atom Description** | **Reporting Rate** | **# Applicable** | **# Filled** | **# Model Reporting Guidelines requesting** |
| --- | --- | --- | --- | --- |
| Specify who funded / supported the study and clarify any conflicts of interest | 0.0% | 10 | 0 | 4 |
| Information on how to access the data used | 0.0% | 12 | 0 | 4 |
| Provide statistics on the amount of missing data there is. | 8.3% | 12 | 1 | 5 |
| Given the problem context, describe what factors or subgroups would be helpful to do a sub-analysis of model performance evaluation (e.g. demographics, environment, lighting)? These factors do not have to be available in the data. | 41.7% | 12 | 5 | 5 |
| Provide summary statistics of key demographics/characteristics/other factors for the data set in question | 50.0% | 12 | 6 | 6 |
| Age is an important demographic factor to report summary statistics on or disaggregate performance by | 50.0% | 12 | 6 | 4 |
| Sex is an important demographic factor to report summary statistics on or disaggregate performance by | 33.3% | 12 | 4 | 4 |
| Other factors for the prediction problem are important to discussed to report summary statistics on or disaggregate performance by (e.g. Gender, Sexual orientation, Fitzpatrick skin type, Socioeconomic Status, Geographic Location, Presenting Symptoms/Clinical Signs/Lab Values/Other Diagnoses) | 50.0% | 12 | 6 | 4 |
| Flow chart of how participants were interacted/assigned/followed up with in the study (especially in clinical trials) | 0.0% | 12 | 0 | 5 |
| Describe the annotation process of the input data, including who annotated the input data, what instructions they were given, and what expertise was needed. | 18.2% | 11 | 2 | 4 |
| Blinding of Data Collectors/Predictor Assessors to outcomes, if done | 0.0% | 9 | 0 | 4 |
| Describe the annotation process of the output data, including who annotated the output data, what instructions they were given, and what expertise was needed. | 27.3% | 11 | 3 | 7 |
| Blinding of Outcome Assessors to predictors of the model, if done | 0.0% | 9 | 0 | 7 |
| How missing data were handled | 50.0% | 12 | 6 | 10 |
| If feature selection involved computing univariate associations between input features and outcomes (not recommended), document this. | 18.2% | 11 | 2 | 4 |
| Provide confidence intervals, statistical significance, or some other handling of uncertainty and variability in model performance metrics | 0.0% | 12 | 0 | 10 |
| Provide sufficient information to enable reproducibility/replication | 0.0% | 12 | 0 | 7 |
| Report model coefficients (regression) or saliency map | 8.3% | 12 | 1 | 7 |
| Disaggregate performance by subgroup / other important data slice | 33.3% | 12 | 4 | 8 |
| Describe external validation strategy / evaluation data set, e.g. what external data set was used, ways it may differ from the training set (e.g. geography, time), why the data set was chosen | 33.3% | 12 | 4 | 9 |
| Calibration Plot | 0.0% | 12 | 0 | 6 |
| NPV | 16.7% | 12 | 2 | 6 |
| Sensitivity, ideally at a predefined probability threshold. | 41.7% | 12 | 5 | 9 |
| Specificity, ideally at a predefined probability threshold. | 8.3% | 12 | 1 | 8 |
| Net Reclassification Improvement | 0.0% | 12 | 0 | 5 |
| Specify what directions, explanations and other user-facing materials there will be with the model. | 0.0% | 12 | 0 | 9 |
| Guidance on how to deploy ML model into clinical workflows | 33.3% | 12 | 4 | 7 |
| Which version of the model is discussed | 45.5% | 11 | 5 | 6 |
| Describe how models are updated/locally tuned | 41.7% | 12 | 5 | 8 |

There was low availability of information on missingness-related atoms, including statistics on amount of missing data (8.3%) and how missing data were handled (50%). There was low information on atoms related to interpreting the model and its performance, such as model coefficients (8.3%), confidence intervals or statistical significance in model performance metrics (0%), and performance of an external validation (33%). There was low reporting of guidance on how to deploy the ML model into a clinical workflow (33%), what user-facing materials there will be with the model (0%), and how models are updated (42%). Lastly, some logistical information had 0% completion, including who funded the study (which might be relevant for conflict of interest purposes) and how to access the data set.

## Discussion

This work is one of the first to systematically compile atoms from reporting guidelines and analyze deployed models’ adherence to existing model reporting guidelines. The 220 atoms, compiled from 15 model reporting guidelines, demonstrate the breadth of details that model developers and researchers consider important to report about a model that will guide care. These atoms cover a range of steps in bringing a model into clinical use (Figure 1). Some categories of model development and deployment have many corresponding atoms, while others have few. For example, while there are 28 atoms on model performance metrics, there are few related to deployment design such as work capacity and resources to perform interventions,^7^ and utility assessment, including eliciting stakeholder preferences.^67^

Model Briefs had excellent reporting of the most commonly requested atoms (Table 2): 9 of the 12 most commonly requested atoms had reporting rates above 90%. These included information on model development and use, such as the outcome definition, and how the model is intended to be used.

However, Model Briefs had low completion rates of all applicable atoms (median 39%). We acknowledge that some reporting guidelines were published after some Model Briefs were created, so it may not be reasonable to expect Model Briefs to adhere fully to those reporting guidelines. Nevertheless, the low completion rate overall suggests that the combined request of all atoms may be formidable for model developers to report and adhere to.

Different reporting guidelines have different focus areas in terms of the different stages in creation and evaluation of a machine learning model (Figure 1). Individual reporting guidelines have generally low adherence rates (median 53%), suggesting that it may be infeasible to report everything that the 15 guidelines collectively request. We recommend model developers select the appropriate reporting guidelines based on their focus of interest (Figure 1); e.g. for model development, use TRIPOD; for fairness, use Model Cards.

Lastly, there are many atoms requested by the reporting guidelines, that are not reported in any of the reviewed Model Briefs (Table 4). Broadly, these relate to fairness, utility, reliability and transparency. For atoms relating to fairness (in this case, referring to data set representativeness and model performance for subgroups), there was low reporting of summary statistics or disaggregated performance for race/ethnicity (33%), age (50%), sex (33%), and other relevant factors (50%). Subgroup and intersectional analyses were rarely performed (33%, 0%), despite evidence of algorithms’ discriminatory behavior against individuals in subgroups^2^ and intersectional subgroups.^68^ We further acknowledge this is a limited view of “fairness” (which has an entire dedicated field of scholarship^69^) and that atoms must be contextualized depending on how the model is used and how the data is collected. For example, biased outcome measurement would not be captured by subgroup analyses of performance.^6^

For atoms relating to utility (referring to the net benefit of model use, including from the standpoint of stakeholder values and resource constraints^7,70–76^), none of the Model Briefs reported any utility-related metrics, including the Net Benefit.^32,33,65^ Work capacity^7^ (resources required to perform interventions) or stakeholder preferences^67,77^ were not formally requested by any model reporting guideline, nor reported by any Model Brief. This is despite studies showing that utility-maximizing models may be different from discrimination-maximizing models^78^ and that work capacity must be taken into consideration for models to create net benefit for patients.^7^ Finally, while there was 100% reporting of atoms on both the intended user and intended use of the model in a specific clinical context, more detailed information on deployment was often missing, like specific guidance on how to deploy into a workflow (33%), specific directions or other user-facing material (0%), time of model prediction (33%), and warnings on out-of-scope use (42%) and when to stop use (8.3%).

For atoms relating to reliability (referring to the stable performance of clinical predictive models across time and deployment sites), there was low reporting of atoms regarding missingness, validation, and monitoring. For missingness, missing data statistics and strategy of missingness handling had low reporting rates (8.3% and 50%). For validation, external validation strategy (33%), calibration plots (0%), and performance comparison against a baseline model (58%) also had low reporting. For monitoring, how models are updated and tuned had a low reporting rate of 42%, and other key atoms for monitoring had reporting rates less than 10%, such as monitoring input data (10%) or regressions in prediction quality in newer data (8.3%).

Lastly, on transparency, there was low reporting of information to enable model reproducibility (0%), model coefficients (8.3%), how to access the data set (0%) (acknowledging necessary limits to protect patient privacy), and who funded the study (0%), which might be relevant for conflict of interest purposes. Model Briefs are not accessible to those without an Epic institutional license, which may further hamper reproducibility and independent validation. A recent independent validation of the Epic Sepsis Model indeed found decreased calibration and discrimination.^23^

Low adherence rates when considering entire model reporting guidelines suggest opportunities to better operationalize reporting practices to ensure deployed models are useful, reliable and fair. One might choose among the many available reporting guidelines by tracking which models have reported atoms from which guideline. Such usage analysis would allow prioritization of more relevant and feasible reporting practices. Similarly, we could incentivise improved reporting if models that have better reporting result in higher adoption, perhaps via endorsement from professional societies in a manner similar to clinical practice guidelines. This could be enabled by a public dashboard tracking models’ guideline adherence. Lastly, deployment teams can benefit from adherence to reporting guidelines by using the atoms from them as checklists for assessing usefulness, workflow capacity, reliability monitoring,^27^ and reviewing them at project initiation time.^79^

There are several key limitations of our methods. First of all, our deduplication of the reporting guidelines may mask certain differences -- e.g. some guidelines provide explicit instructions and examples while others just call for reporting. We also caution against over-interpreting the completion rate across all atoms, as atoms are not exchangeable entities. Two atoms such as “Missing data statistics” and “Sensitivity” provide different information, so we recommend looking at individual atoms when possible. Lastly, to provide an upper bound on the quality of reporting, we gave generous credit to Model Briefs for reporting of an atom. For example, we gave credit for “Describe how models were tested in a new setting before deployment” for statements that might have simply stated to contact a support representative to validate the model. Hence reporting rates should be viewed as likely overestimates.

## Conclusion

Despite ongoing discussion on what should be reported about predictive models, adherence of current documentation for deployed models to existing reporting guidelines has not been assessed. In this work, we compiled reportable items from existing reporting guidelines into a set of unique “atoms” and reviewed the documentation of the 12 most adopted models from a widely used health vendor, Epic. We identified 220 distinct atoms, of which 176 were applicable to at least one model.

Current model documentation reports information for less than half of applicable atoms (median 39% per Model Brief), and model reporting guidelines have low adherence rates based on the documentation (median 54% per guideline). Current model documentation provides relatively little information on usefulness, reliability, transparency and fairness. There is a need for better operationalization of reporting practices for predictive models in healthcare.

## Supporting information

eFile 1

## Data Availability

All data and code used for methods, including merging of guidelines, deduplication of atoms, mapping of atoms onto stages of model development and tasks, grading of Model Briefs, adjudication, and analysis are available at eFile 1.

## CONTRIBUTORS

NHS conceived the study. AC, KEM, BSP and NHS provided guidance on experimental design, implementation, analysis and writing for JHL. BSP and JHL searched for the model reporting guidelines. JHL performed deduplication of model reporting guidelines. JHL designed instructions for grading Model Briefs and adjudication. KEM, BSP, AC and JHL independently graded Model Briefs. DD adjudicated disagreements. JHL generated analysis and resulting figures. All authors participated in writing of the final manuscript.

## Acknowledgements

The authors would like to thank Scott Fleming, Siyun Li, Arjun Gokhale, Wui Ip, Lillian Sung, and Ron Li for providing project feedback and guidance, and members of the Shah lab and the Data Science Team at IAT for ideation, moral support, and feedback during a once-in-a-lifetime pandemic. Thanks to Stephen Pfohl for reader feedback.

