## Supplement for "Low adherence to existing model reporting guidelines by commonly used clinical prediction models"

### **eMethods**

In reviewing, reviewers could designate atoms as “reported”, “not reported”, “not applicable” and also “wrongly reported.” However, for “wrongly reported”, this was rare to occur and only one atom for one Model Brief was adjudicated to this, so this is not further discussed in the manuscript.

### **eFIGURES**

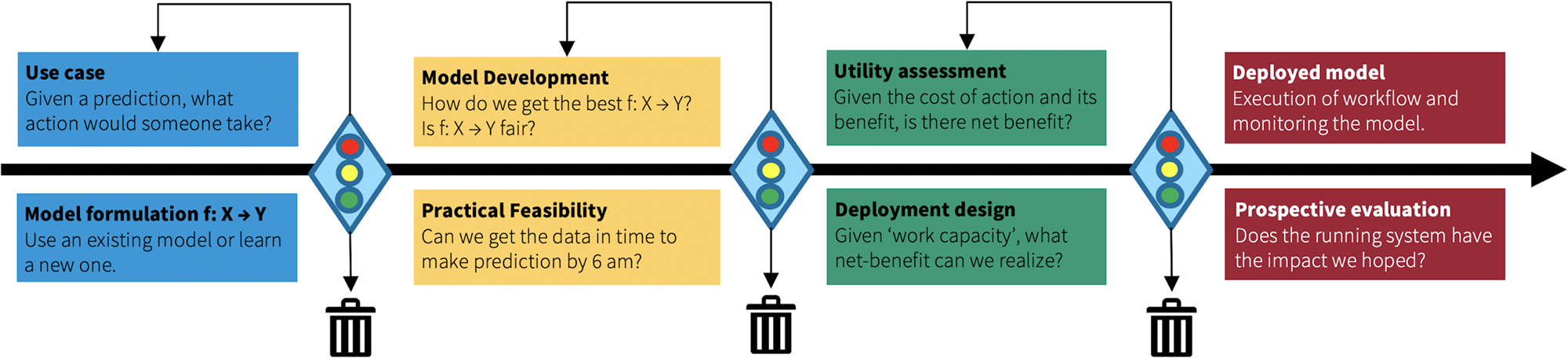

#### **eFigure 1**. Figure 4 reproduced from ^1^, with permission under the Creative Commons CC-BY-NC license. These prompts were used for our atom mapping onto the different stages in the creation and evaluation of a machine learning model to guide care (eFile 1, Figure 1).

### **eTABLES**

|  | **MODEL REPORTING GUIDELINES** | | | | | | | | | | | | | | | **Total Atoms** |
| --- | --- | --- | --- | --- | --- | --- | --- | --- | --- | --- | --- | --- | --- | --- | --- | --- |
| **TASK** | **Model Cards** | **Model Facts Labels** | **Guidelines** | **MI-CLAIM** | **MINIMAR** | **TRIPOD** | **CONSORT-AI** | **SPIRIT-AI** | **Trust and Value** | **ML Test Score** | **Risk** | **STARD** | **ABCD** | **CHARMS** | **PROBAST** |  |
| **Overview** | 7 | 8 | 6 | 2 | 1 | 10 | 9 | **14** | 1 | 0 | 1 | 8 | 2 | 2 | 2 | 28 |
| Overview: Clinical Trial | 0 | 1 | 0 | 0 | 0 | 1 | 2 | **9** | 0 | 0 | 0 | 2 | 0 | 0 | 0 | 9 |
| **Data Composition** | 7 | 4 | 8 | 6 | 9 | 10 | 8 | 4 | 1 | 3 | 3 | 10 | 5 | 10 | 11 | 24 |
| Data Composition: Input | 0 | 1 | 1 | 1 | 1 | 3 | 0 | 0 | 0 | 0 | 1 | 0 | 1 | 3 | **4** | 5 |
| Data Composition: Factors | **6** | 0 | 0 | 2 | 5 | 1 | 5 | 0 | 0 | 3 | 0 | 5 | 0 | 0 | 0 | 7 |
| Data Composition: Output | 0 | 2 | 3 | 1 | 1 | 2 | 1 | 1 | 0 | 0 | 1 | 2 | 3 | **4** | **4** | 7 |
| **Study Design/Population** | 1 | 2 | 2 | 0 | 3 | **4** | **4** | **4** | 0 | 0 | 2 | 2 | 1 | 3 | **4** | 4 |
| **Data Collection & Methods** | 6 | 1 | 1 | 0 | 0 | 11 | 4 | **12** | 6 | 1 | 5 | 8 | 0 | 9 | 10 | 21 |
| Data Collection & Methods: Input | 1 | 0 | 0 | 0 | 0 | **2** | 1 | 1 | 0 | 0 | 0 | 0 | 0 | 1 | **2** | 2 |
| Data Collection & Methods: Outcome | 1 | 0 | 0 | 0 | 0 | 4 | 1 | 2 | 1 | 0 | 2 | 4 | 0 | 4 | **5** | 7 |
| Data Collection & Methods: Consent/Privacy for Data | 0 | 0 | 0 | 0 | 0 | 0 | 0 | **4** | **4** | 1 | 0 | 0 | 0 | 0 | 0 | 4 |
| Evaluation-specific Study Details: Methods | 0 | 0 | 0 | 1 | 0 | 2 | 5 | **10** | 1 | 0 | 2 | 0 | 0 | 3 | 0 | 13 |
| Evaluation-specific Study Details: Randomization | 0 | 0 | 0 | 0 | 0 | 0 | **4** | 3 | 0 | 0 | 1 | 0 | 0 | 0 | 0 | 4 |
| Evaluation-specific Study Details: Blinding | 0 | 0 | 0 | 0 | 0 | 0 | 3 | **5** | 0 | 0 | 0 | 0 | 0 | 0 | 0 | 5 |
| Evaluation-specific Study Details: Outcomes | 0 | 0 | 0 | 0 | 0 | 0 | **4** | 1 | 0 | 0 | 1 | 0 | 0 | 0 | 0 | 4 |
| Evaluation-specific Study Details: Analysis | 0 | 1 | 0 | 0 | 0 | 0 | **4** | **4** | 1 | 0 | 2 | 0 | 0 | 1 | 0 | 4 |
| **Preprocessing and Data Cleaning** | 1 | 0 | 3 | 1 | 2 | 4 | 3 | 2 | 0 | 1 | 5 | 1 | 4 | 5 | **6** | 7 |
| **Model Building** | 1 | 0 | 2 | 3 | 2 | **5** | 1 | 1 | 0 | 3 | 3 | 1 | 3 | 3 | 2 | 8 |
| **Model Summary** | 2 | 1 | 1 | 1 | 1 | **3** | 1 | 1 | 0 | 0 | 2 | 2 | 1 | **3** | 1 | 4 |
| **Model Performance and Comparison** | **1** | 0 | **1** | **1** | 0 | **1** | **1** | 0 | **1** | 0 | **1** | **1** | **1** | **1** | 0 | 1 |
| **Model Examination** | 2 | 0 | 5 | **9** | 0 | 1 | 4 | 3 | 1 | 2 | 1 | 1 | 2 | 3 | 1 | 13 |
| **Validation** | 3 | 3 | 2 | 2 | 3 | **4** | 0 | 0 | 3 | 0 | 3 | 0 | 3 | 3 | 3 | 4 |
| **Metrics** | 7 | 4 | 8 | 8 | 5 | **17** | 0 | 0 | 1 | 1 | 5 | 12 | 10 | 10 | 10 | 29 |
| Metrics: Discrimination | 1 | 1 | 1 | 1 | 1 | **3** | 0 | 0 | 0 | 0 | 1 | 1 | 2 | 1 | 1 | 3 |
| Metrics: Goodness-of-Fit | 0 | 0 | 1 | 0 | 0 | **3** | 0 | 0 | 0 | 0 | 0 | 1 | 1 | 2 | 1 | 6 |
| Metrics: Calibration | 0 | 0 | 1 | 0 | 0 | **2** | 0 | 0 | 0 | 0 | 1 | 0 | **2** | 1 | 1 | 2 |
| Metrics: Classification | 5 | 2 | 4 | 5 | 3 | 4 | 0 | 0 | 0 | 0 | 0 | **9** | 2 | 4 | 4 | 12 |
| Metrics: Utility | 0 | 0 | 0 | 1 | 0 | **2** | 0 | 0 | 0 | 0 | 0 | 0 | 1 | 0 | 1 | 3 |
| Metrics: Compare Two Model Discrimination | 0 | 0 | 0 | 0 | 0 | **2** | 0 | 0 | 0 | 0 | **2** | 0 | 1 | 1 | 1 | 2 |
| **Comparison Against Baseline Model** | 0 | 0 | 2 | **3** | 0 | 2 | 0 | 0 | 0 | 2 | 0 | 1 | 0 | 0 | 0 | 3 |
| **Intended Use** | 3 | **6** | 3 | 1 | 2 | 3 | 3 | 3 | 3 | 0 | 2 | 3 | 1 | 1 | 1 | 7 |
| Intended Use: User | 1 | **2** | 1 | 0 | 1 | 1 | **2** | **2** | 1 | 0 | 1 | 1 | 1 | 0 | 0 | 2 |
| Intended Use: Warnings | 1 | **2** | 0 | 0 | 0 | 0 | 0 | 0 | 1 | 0 | 0 | 0 | 0 | 0 | 0 | 3 |
| **Deployment** | 2 | 5 | 1 | 1 | 0 | 4 | 5 | 6 | 6 | **20** | 2 | 1 | 0 | 3 | 2 | 26 |
| Deployment: Tests: Data | 1 | 0 | 0 | 0 | 0 | 0 | 0 | 1 | 1 | **5** | 0 | 0 | 0 | 0 | 0 | 6 |
| Deployment: Tests: Infrastructure | 0 | 0 | 0 | 0 | 0 | 0 | 0 | 0 | 0 | **4** | 0 | 0 | 0 | 0 | 0 | 4 |
| Deployment: Updating | 1 | 2 | 0 | 0 | 0 | **4** | 3 | 3 | 2 | **4** | 2 | 0 | 0 | 2 | 1 | 6 |
| Deployment: Monitoring | 0 | 1 | 1 | 0 | 0 | 0 | 0 | 0 | 1 | **7** | 0 | 0 | 0 | 0 | 0 | 7 |
| **Ethics** | **3** | 1 | 1 | 0 | 0 | 0 | 1 | 1 | 1 | 0 | 0 | 1 | 0 | 0 | 0 | 3 |
| **Limitations** | 2 | 1 | 3 | 1 | 0 | **4** | **4** | 1 | 0 | 0 | 0 | 3 | 0 | 2 | 1 | 6 |
| Miscellaneous | **1** | 0 | 0 | 0 | 0 | **1** | 0 | 0 | 0 | **1** | 0 | 0 | 0 | **1** | **1** | 2 |

#### **eTable 1: Model Reporting Guidelines by Tasks.** Model reporting guidelines (in rows), with their atoms mapped onto different tasks in model development and deployment. The highest number in each row is bolded. Cells are shaded if they provide less than half of the total atoms.

| **Atom Description** | **# Model Reporting Guidelines requesting** | **Task** | **Stage** | **Reporting Rate** |
| --- | --- | --- | --- | --- |
| AUROC (c- index) | 11 | Metrics: Discrimination | Model Development | 91.67% |
| Prognostic Index Plot for Validation Data Set | 1 | Metrics: Discrimination | Model Development | 0.00% |
| Any direct examination of model output, for example a Plot to Visualize Discrimination | 2 | Metrics: Discrimination | Model Development | 83.33% |
| Normalized root-mean squared error | 1 | Metrics: Goodness-of-Fit | Model Development | 16.67% |
| R^2 | 1 | Metrics: Goodness-of-Fit | Model Development | 8.33% |
| Brier Score | 1 | Metrics: Goodness-of-Fit | Model Development | 0.00% |
| D-statistic | 3 | Metrics: Goodness-of-Fit | Model Development | 0.00% |
| For survival curves, the log-rank test | 1 | Metrics: Goodness-of-Fit | Model Development | N/A |
| Odds Ratio of two different models for comparison | 2 | Metrics: Goodness-of-Fit | Model Development | 0.00% |
| Calibration Plot | 6 | Metrics: Calibration | Model Development | 0.00% |
| Survival Curve/Kaplan-Meier Curve superimposition (for Cox models) | 2 | Metrics: Calibration | Model Development | N/A |
| PPV | 8 | Metrics: Classification | Model Development | 66.67% |
| NPV | 6 | Metrics: Classification | Model Development | 16.67% |
| Sensitivity, ideally at a predefined probability threshold. | 9 | Metrics: Classification | Model Development | 41.67% |
| Specificity, ideally at a predefined probability threshold. | 8 | Metrics: Classification | Model Development | 8.33% |
| Full Contingency Table against Reference (includes True/False Positives/Negatives) | 2 | Metrics: Classification | Model Development | 0.00% |
| True Positive (TP) | 1 | Metrics: Classification | Model Development | 8.33% |
| True Negative (TN) | 1 | Metrics: Classification | Model Development | 0.00% |
| False Positive / False Positive Rate | 2 | Metrics: Classification | Model Development | 16.67% |
| False Negative / False Negative Rate | 2 | Metrics: Classification | Model Development | 8.33% |
| False Discovery Rate | 1 | Metrics: Classification | Model Development | 0.00% |
| False Omission Rate | 1 | Metrics: Classification | Model Development | 0.00% |
| F score / Dice Coefficient | 1 | Metrics: Classification | Model Development | 0.00% |
| NNT | 1 | Metrics: Utility | Utility Assessment | 0.00% |
| Net Benefit (Decision Curve) | 3 | Metrics: Utility | Utility Assessment | 0.00% |
| Relative Utility (Decision Curve) | 1 | Metrics: Utility | Utility Assessment | 0.00% |
| Net Reclassification Improvement | 5 | Metrics: Compare Two Model Discrimination | Model Development | 0.00% |
| Integrated Discrimination Improvement | 2 | Metrics: Compare Two Model Discrimination | Model Development | 0.00% |

#### **eTable 2: Requested Metrics**. All atoms requested, relating to a model performance metric are listed. Reporting Rate indicates the % of the Model Briefs that provided the information requested in the atom; N/A means the atom did not apply to any Model Briefs. Task and Stage indicate the atoms’ related task and related stage of clinical predictive model development, respectively ^1^.

| **Atom Description** | **Requesting Model Reporting Guideline** | **Task** | **Stage** | **Reporting Rate** |
| --- | --- | --- | --- | --- |
| How should the model be cited? | Model Cards | Overview | Other: Logistics | 0.00% |
| Is the model regulated or approved by the FDA? | Trust and Value | Overview | Deployed Model: Monitoring | 0.00% |
| Model Name | Model Facts Labels | Overview | Other: Logistics | 100.00% |
| For clinical trials, names and roles of folks involved in clinical trial protocol. | SPIRIT-AI | Overview: Clinical Trial | Other: Logistics | N/A |
| For clinical trials, names and roles of individuals/groups who oversee. | SPIRIT-AI | Overview: Clinical Trial | Other: Personnel | N/A |
| For clinical trials, sponsor contact info | SPIRIT-AI | Overview: Clinical Trial | Other: Personnel | N/A |
| For clinical trials, plans/status of research ethics review approval. | SPIRIT-AI | Overview: Clinical Trial | Other: Logistics | N/A |
| For clinical trials, plan to communicate or report results to participants and other stakeholders | SPIRIT-AI | Overview: Clinical Trial | Other: Logistics | N/A |
| For clinical trials, Authorship eligibility guidelines and any intended use of professional writers. | SPIRIT-AI | Overview: Clinical Trial | Other: Personnel | N/A |
| For clinical trials, Plans for collection, laboratory evaluation, and storage of biological specimens for genetic or molecular analysis in the current trial and for future use in ancillary studies, if applicable. | SPIRIT-AI | Overview: Clinical Trial | Other: Logistics | N/A |
| It is clear what each data point (i.e. what does a n=1 mean?) of the data set is. | Guidelines | Data Composition | Model Development | 100.00% |
| Clarify if input data is structured (defined like medications) or unstructured (pixels, natural language, time series) | MI-CLAIM | Data Composition: Input | Model Formulation | 100.00% |
| Has there been a check on input features that correlate with protected user categories, which may lead to uninclusive, privacy-breaching or discriminatory results? | ML Test Score | Data Composition: Factors | Model Development: Fairness | 8.33% |
| Report the distribution of severity/stage of disease in those with the target condition | STARD | Data Composition: Output | Model Development | 0.00% |
| Report the distribution of alternative diagnoses in those without the target condition. | STARD | Data Composition: Output | Model Development | 8.33% |
| Describe if data annotators were given compensation. | Model Cards | Data Collection & Methods | Other: Personnel | 0.00% |
| If there was a time interval and any interventions that occurred between the diagnostic index test and the reference standard, report it. | STARD | Data Collection & Methods: Outcome | Model Development | 57.14% |
| The time interval between the assessment of the predictors and the outcome is appropriate to allow the correct type and representative number of relevant outcomes to be recorded | PROBAST | Data Collection & Methods: Outcome | Model Development | 91.67% |
| Define any strategies for improving and monitoring adherence to interventions. | SPIRIT-AI | Evaluation-specific Study Details: Methods | Prospective Evaluation | N/A |
| Clarify any retention and follow-up strategies for patients. | SPIRIT-AI | Evaluation-specific Study Details: Methods | Prospective Evaluation | N/A |
| Plan to communicate clinical protocol amendments to all relevant stakeholders | SPIRIT-AI | Evaluation-specific Study Details: Methods | Other: Logistics | N/A |
| Changes to clinical trial methods after start of trial | CONSORT-AI | Evaluation-specific Study Details: Methods | Prospective Evaluation | N/A |
| Guidelines for when / why to stop clinical trial | CONSORT-AI | Evaluation-specific Study Details: Methods | Prospective Evaluation | N/A |
| Why trial ended or was stopped | CONSORT-AI | Evaluation-specific Study Details: Methods | Prospective Evaluation | N/A |
| For clinical trials, structure and role of the clinical trial data monitoring committee | SPIRIT-AI | Evaluation-specific Study Details: Methods | Other: Personnel | N/A |
| For clinical trials, discuss who audits conduct | SPIRIT-AI | Evaluation-specific Study Details: Methods | Other: Personnel | N/A |
| Ancillary and post-trial care/compensation for those suffering harm | SPIRIT-AI | Evaluation-specific Study Details: Methods | Other: Logistics | N/A |
| Details on how similar interventions are and relation to concealment | SPIRIT-AI | Evaluation-specific Study Details: Blinding | Prospective Evaluation | N/A |
| Emergency unblinding procedures | SPIRIT-AI | Evaluation-specific Study Details: Blinding | Prospective Evaluation | N/A |
| Clinical Outcomes: any changes to definition | CONSORT-AI | Evaluation-specific Study Details: Outcomes | Prospective Evaluation | N/A |
| Define the results of the clinical outcomes based on definitions. | CONSORT-AI | Evaluation-specific Study Details: Outcomes | Prospective Evaluation | 100.00% |
| Clinical Outcomes: Binary Outcomes show both absolute and relative effect sizes | CONSORT-AI | Evaluation-specific Study Details: Outcomes | Prospective Evaluation | N/A |
| Provide a check that model training is reproducible | ML Test Score | Model Building | Model Development | 8.33% |
| Check that training/learning objectives for ML are correlated with desired clinical impact metrics | ML Test Score | Model Building | Model Development | 0.00% |
| How indeterminate model outputs were handled | STARD | Model Summary | Model Formulation | 0.00% |
| Report most predictive features of model | Guidelines | Model Examination | Model Development | 8.33% |
| A check to see if each feature is helping predictive power | ML Test Score | Model Examination | Model Development | 58.33% |
| Disaggregate performance by intersection of subgroups | Model Cards | Model Examination | Model Development: Fairness | 0.00% |
| Report at least 2 distinct model examinations. | MI-CLAIM | Model Examination | Model Development | 100.00% |
| Perform a sensitivity analysis of the model | MI-CLAIM | Model Examination | Model Development | 8.33% |
| Discuss the model examination and performance tradeoffs | MI-CLAIM | Model Examination | Model Development | 25.00% |
| Discuss model reliability under distribution shifts. | MI-CLAIM | Model Examination | Model Development | 8.33% |
| Describe how predictions were calculated in an external validation | TRIPOD | Validation | Model Development | 9.09% |
| Prognostic Index Plot for Validation Data Set | TRIPOD | Metrics: Discrimination | Model Development | 0.00% |
| Normalized root-mean squared error | Guidelines | Metrics: Goodness-of-Fit | Model Development | 16.67% |
| R^2 | TRIPOD | Metrics: Goodness-of-Fit | Model Development | 8.33% |
| Brier Score | TRIPOD | Metrics: Goodness-of-Fit | Model Development | 0.00% |
| For survival curves, the log-rank test | CHARMS | Metrics: Goodness-of-Fit | Model Development | N/A |
| True Positive (TP) | STARD | Metrics: Classification | Model Development | 8.33% |
| True Negative (TN) | STARD | Metrics: Classification | Model Development | 0.00% |
| False Discovery Rate | Model Cards | Metrics: Classification | Model Development | 0.00% |
| False Omission Rate | Model Cards | Metrics: Classification | Model Development | 0.00% |
| F score / Dice Coefficient | MI-CLAIM | Metrics: Classification | Model Development | 0.00% |
| NNT | MI-CLAIM | Metrics: Utility | Utility Assessment | 0.00% |
| Relative Utility (Decision Curve) | TRIPOD | Metrics: Utility | Utility Assessment | 0.00% |
| Description of the clinical activity or annoyance that may be required to make the model, e.g. manually enter info, move to another screen, or otherwise make additional "clicks"? | Trust and Value | Intended Use: Warnings | Use Case | 8.33% |
| A warning on when to stop use of model | Model Facts Labels | Intended Use: Warnings | Use Case | 8.33% |
| Check that no feature costs too much to have (e.g. dependencies, latency, instability, maintenance costs) compared with its added predictive value | ML Test Score | Deployment: Tests: Data | Practical Feasibility | 0.00% |
| Programmatically enforce that input features adhere to meta-level requirements (e.g. deprecated features, protected features). | ML Test Score | Deployment: Tests: Data | Deployed Model: Execution | 0.00% |
| In deployment, a new feature can be added quickly (e.g. within 1-2 months) to the model from ideation. | ML Test Score | Deployment: Tests: Data | Deployed Model: Execution | 0.00% |
| Develop unit tests for input features. | ML Test Score | Deployment: Tests: Data | Deployed Model: Monitoring | 0.00% |
| Monitor input data to ensure that it falls within correct ranges and invariances. | ML Test Score | Deployment: Tests: Data | Deployed Model: Monitoring | 0.00% |
| Perform unit tests of model specification: API usage (e.g. check API calls on a random input) and algorithmic correctness (is it producing the predictions for the correct reasons)? | ML Test Score | Deployment: Tests: Infrastructure | Deployed Model: Monitoring | 0.00% |
| Continuously perform an integration test: a fully automated test that runs regularly and uses the entire pipeline, validating that data and code can successfully move through each stage and that the resulting model performs well. | ML Test Score | Deployment: Tests: Infrastructure | Deployed Model: Monitoring | 0.00% |
| Model allows debugging by step-by-step computation of training/inference on single example | ML Test Score | Deployment: Tests: Infrastructure | Deployed Model: Monitoring | 0.00% |
| Models are tested via a canary process before they enter production serving environments: | ML Test Score | Deployment: Tests: Infrastructure | Deployed Model: Monitoring | 0.00% |
| Report the parts of the models that have been updated and the performance of the updated model | TRIPOD | Deployment: Updating | Deployed Model: Monitoring | 16.67% |
| Every model specification undergoes a code review and is checked in to a repository: | ML Test Score | Deployment: Updating | Deployed Model: Monitoring | 0.00% |
| Check that models can be quickly, easily rolled back in case of emergency. | ML Test Score | Deployment: Updating | Deployed Model: Monitoring | 0.00% |
| Monitor the age of the model and determine how old will affect the staleness of the model. | ML Test Score | Deployment: Monitoring | Deployed Model: Monitoring | 0.00% |
| The deployment team has a line of communication with upstream, dependent data sources and is familiar with new data source changes. | ML Test Score | Deployment: Monitoring | Deployed Model: Monitoring | 0.00% |
| Programmatically check whether data matches invariants in schema and alert when they diverge significantly, tuning a reasonable false positive/false negative point. | ML Test Score | Deployment: Monitoring | Deployed Model: Monitoring | 0.00% |
| Check that training and serving features compute the same values, either by direct comparison of features computed in both systems, or by comparing distributions. | ML Test Score | Deployment: Monitoring | Deployed Model: Monitoring | 0.00% |
| Check degradations in the model computational performance. | ML Test Score | Deployment: Monitoring | Deployed Model: Monitoring | 0.00% |
| Discuss any risk mitigation strategies used during model development. | Model Cards | Ethics | Model Development: Fairness | 25.00% |
| Acknowledge if the model is intended to inform decisions about human life or safety. | Model Cards | Ethics | Use Case | 91.67% |
| Describe any pitfalls in interpreting the model. | Guidelines | Limitations | Model Development | 33.33% |

#### **eTable 3: Uniquely Requested Atoms**. All atoms requested by exactly 1 model reporting guideline are listed. Reporting Rate indicates the % of the Model Briefs that provided the information requested in the atom. Task and Stage indicate the atoms’ related task and related stage of clinical predictive model development, respectively ^1^.

| **Atom Description** | **Reporting Rate** | **# Applicable** | **# Filled** | **# Model Reporting Guidelines requesting** | **Task** | **Stage** |
| --- | --- | --- | --- | --- | --- | --- |
| Who and how to contact with questions about the model | 100.00% | 12 | 12 | 2 | Overview | Other: Personnel |
| Model Name | 100.00% | 12 | 12 | 1 | Overview | Other: Logistics |
| Date of model development and/or last update | 100.00% | 12 | 12 | 2 | Overview | Model Formulation |
| Model one-line summary | 100.00% | 12 | 12 | 2 | Overview | Model Formulation |
| Scientific / clinical background and rationale for model use (e.g. previous work, clinical role) | 100.00% | 12 | 12 | 6 | Overview | Use Case |
| Specify the type of prediction problem: classification, regression, survival prediction | 91.67% | 12 | 11 | 2 | Overview | Model Formulation |
| Specify whether the data/study was retrospective or prospective. | 100.00% | 12 | 12 | 3 | Overview | Model Development |
| Specify whether the data/study was prognostic or diagnostic? | 100.00% | 12 | 12 | 4 | Overview | Model Formulation |
| Summarize, discuss and interpret results | 91.67% | 12 | 11 | 2 | Overview | Other |
| Specify who (person/organization) built the model | 100.00% | 12 | 12 | 2 | Overview | Other: Personnel |
| Provide any description of the data set (training / study) in question | 100.00% | 12 | 12 | 12 | Data Composition | Model Development |
| For the data set in question, what the sample size is and how it was arrived at, if pre-specified (e.g. Events Per Variable minima) | 91.67% | 12 | 11 | 9 | Data Composition | Model Development |
| It is clear what each data point (i.e. what does a n=1 mean?) of the data set is. | 100.00% | 12 | 12 | 1 | Data Composition | Model Development |
| Describe, list and/or define all input features | 100.00% | 12 | 12 | 7 | Data Composition: Input | Model Formulation |
| Clarify if input data is structured (defined like medications) or unstructured (pixels, natural language, time series) | 100.00% | 12 | 12 | 1 | Data Composition: Input | Model Formulation |
| It is clear if there is a reasonable number of Events per predictor (typically >= 10 or 20)? | 91.67% | 12 | 11 | 4 | Data Composition: Input | Model Development |
| It is clear if candidate predictors are available at time of intended use of model? | 91.67% | 12 | 11 | 2 | Data Composition: Input | Practical Feasibility |
| Define the output/outcome produced by the model | 100.00% | 12 | 12 | 10 | Data Composition: Output | Model Formulation |
| It is clear whether the outcome is a single or combined endpoint (e.g. cardiovascular disease including heart disease and stroke) | 100.00% | 12 | 12 | 2 | Data Composition: Output | Model Development |
| Define the target population of the data in question (who the model should generalize / apply to?) | 100.00% | 12 | 12 | 8 | Study Design/Population | Use Case |
| Define the specific inclusion/exclusion criteria for participants in data (especially in clinical trials) | 100.00% | 12 | 12 | 9 | Study Design/Population | Model Development |
| Define the specific local area/environment/setting of training data / model deployment. | 100.00% | 12 | 12 | 10 | Study Design/Population | Use Case |
| Define the timeline of data collection. This could, for example, include participant recruitment time, time of predictor measurement, and outcome measurement/followup time. | 100.00% | 12 | 12 | 9 | Data Collection & Methods | Model Development |
| Details of treatments received by participants, if relevant. (NOT studying specific interventions for patients, just what treatments they may be receiving already) | 90.91% | 11 | 10 | 2 | Data Collection & Methods | Model Development |
| Consistent Outcome Definition and Measurement for all patients | 100.00% | 12 | 12 | 3 | Data Collection & Methods: Outcome | Model Development |
| Predictors Not Part of Outcome (e.g. in panel or consensus diagnosis) | 100.00% | 12 | 12 | 2 | Data Collection & Methods: Outcome | Model Development |
| The time interval between the assessment of the predictors and the outcome is appropriate to allow the correct type and representative number of relevant outcomes to be recorded | 91.67% | 12 | 11 | 1 | Data Collection & Methods: Outcome | Model Development |
| Define evaluation outcomes for intervention assessment. | 100.00% | 1 | 1 | 3 | Evaluation-specific Study Details: Outcomes | Prospective Evaluation |
| Define the results of the clinical outcomes based on definitions. | 100.00% | 1 | 1 | 1 | Evaluation-specific Study Details: Outcomes | Prospective Evaluation |
| How data was preprocessed (data cleaning, predictor transformation, outlier removal, predictor coding) | 100.00% | 12 | 12 | 10 | Preprocessing and Data Cleaning | Model Development |
| Clarify the type of final model to be used | 91.67% | 12 | 11 | 9 | Model Summary | Model Formulation |
| Report at least 2 distinct model examinations. | 100.00% | 12 | 12 | 1 | Model Examination | Model Development |
| Clarify what type of validation is done, whether internal or external | 100.00% | 12 | 12 | 11 | Validation | Model Development |
| Describe internal validation strategy to account for model optimism (e.g. cross-validation, bootstrapping, data splitting)) | 91.67% | 12 | 11 | 11 | Validation | Model Development |
| Mention what performance measures are used | 100.00% | 12 | 12 | 13 | Metrics | Model Development |
| AUROC (c- index) | 91.67% | 12 | 11 | 11 | Metrics: Discrimination | Model Development |
| Describe how the ML model is supposed to be used in clinical context | 100.00% | 12 | 12 | 11 | Intended Use | Use Case |
| Suggest ways the ML model could impact clinical care (no study needed, it's okay if this is speculative) | 100.00% | 12 | 12 | 6 | Intended Use | Utility Assessment |
| Specify who will use the ML model. | 100.00% | 12 | 12 | 5 | Intended Use: User | Use Case |
| Acknowledge if the model is intended to inform decisions about human life or safety. | 91.67% | 12 | 11 | 1 | Ethics | Use Case |

#### **eTable 4: Commonly Reported Atoms**. All atoms reported by 90% or more of applicable Model Briefs are listed. Reporting Rate indicates the % of the Model Briefs that provided the information requested in the atom. Task and Stage indicate the atoms’ related task and related stage of clinical predictive model development, respectively ^1^.

| **Atom Description** | **Reporting Rate** | **# Applicable** | **# Filled** | **# Model Reporting Guidelines requesting** | **Task** | **Stage** |
| --- | --- | --- | --- | --- | --- | --- |
| How should the model be cited? | 0.00% | 11 | 0 | 1 | Overview | Other: Logistics |
| Is the model regulated or approved by the FDA? | 0.00% | 12 | 0 | 1 | Overview | Deployed Model: Monitoring |
| Specify who funded / supported the study and clarify any conflicts of interest | 0.00% | 10 | 0 | 4 | Overview | Other: Personnel |
| Information on how to access the data used | 0.00% | 12 | 0 | 4 | Data Composition | Other: Logistics |
| Provide statistics on the amount of missing data there is. | 8.33% | 12 | 1 | 5 | Data Composition | Model Development |
| Has there been a check on input features that correlate with protected user categories, which may lead to uninclusive, privacy-breaching or discriminatory results? | 8.33% | 12 | 1 | 1 | Data Composition: Factors | Model Development: Fairness |
| Report the distribution of severity/stage of disease in those with the target condition | 0.00% | 11 | 0 | 1 | Data Composition: Output | Model Development |
| Report the distribution of alternative diagnoses in those without the target condition. | 8.33% | 12 | 1 | 1 | Data Composition: Output | Model Development |
| Flow chart of how participants were interacted/assigned/followed up with in the study (especially in clinical trials) | 0.00% | 12 | 0 | 5 | Data Collection & Methods | Model Development |
| Describe if data annotators were given compensation. | 0.00% | 12 | 0 | 1 | Data Collection & Methods | Other: Personnel |
| Describe the interobserver/inter-study agreement on data coding, and if there was any standardization effort. | 0.00% | 12 | 0 | 3 | Data Collection & Methods | Model Development |
| Blinding of Data Collectors/Predictor Assessors to outcomes, if done | 0.00% | 9 | 0 | 4 | Data Collection & Methods: Input | Model Development |
| Blinding of Outcome Assessors to predictors of the model, if done | 0.00% | 9 | 0 | 7 | Data Collection & Methods: Outcome | Model Development |
| Define who obtains consent to data collection | 0.00% | 12 | 0 | 2 | Data Collection & Methods: Consent/Privacy for Data | Other: Personnel |
| Define what form or measures are taken to ensure informed consent for patients. | 0.00% | 12 | 0 | 2 | Data Collection & Methods: Consent/Privacy for Data | Other: Logistics |
| Define what provisions are taken for participant data use in followup studies. | 0.00% | 12 | 0 | 2 | Data Collection & Methods: Consent/Privacy for Data | Other: Logistics |
| Define how confidentiality and privacy will be ensured for participants' data. | 0.00% | 12 | 0 | 3 | Data Collection & Methods: Consent/Privacy for Data | Other: Logistics |
| Provide a check that model training is reproducible | 8.33% | 12 | 1 | 1 | Model Development | Model Development |
| Check that training/learning objectives for ML are correlated with desired clinical impact metrics | 0.00% | 12 | 0 | 1 | Model Development | Model Development |
| Describe which features were allowed interactions. | 0.00% | 12 | 0 | 2 | Model Development | Model Development |
| Provide confidence intervals, statistical significance, or some other handling of uncertainty and variability in model performance metrics | 0.00% | 12 | 0 | 10 | Model Performance and Comparison | Model Development |
| Provide sufficient information to enable reproducibility/replication | 0.00% | 12 | 0 | 7 | Model Development | Other: Logistics |
| How indeterminate model outputs were handled | 0.00% | 12 | 0 | 1 | Model Summary | Model Formulation |
| Report model coefficients (regression) or saliency map | 8.33% | 12 | 1 | 7 | Model Examination | Model Development |
| Report most predictive features of model | 8.33% | 12 | 1 | 1 | Model Examination | Model Development |
| Describe cases where the model had high or low performance error | 8.33% | 12 | 1 | 2 | Model Examination | Model Development |
| Disaggregate performance by intersection of subgroups | 0.00% | 12 | 0 | 1 | Model Examination | Model Development: Fairness |
| Perform a sensitivity analysis of the model | 8.33% | 12 | 1 | 1 | Model Examination | Model Development |
| Analyze the model's performance errors | 0.00% | 12 | 0 | 2 | Model Examination | Model Development |
| Discuss model reliability under distribution shifts. | 8.33% | 12 | 1 | 1 | Model Examination | Model Development |
| Describe how predictions were calculated in an external validation | 9.09% | 11 | 1 | 1 | Validation | Model Development |
| Prognostic Index Plot for Validation Data Set | 0.00% | 10 | 0 | 1 | Metrics: Discrimination | Model Development |
| R^2 | 8.33% | 12 | 1 | 1 | Metrics: Goodness-of-Fit | Model Development |
| Brier Score | 0.00% | 12 | 0 | 1 | Metrics: Goodness-of-Fit | Model Development |
| D-statistic | 0.00% | 1 | 0 | 3 | Metrics: Goodness-of-Fit | Model Development |
| Odds Ratio of two different models for comparison | 0.00% | 12 | 0 | 2 | Metrics: Goodness-of-Fit | Model Development |
| Calibration Plot | 0.00% | 12 | 0 | 6 | Metrics: Calibration | Model Development |
| Specificity, ideally at a predefined probability threshold. | 8.33% | 12 | 1 | 8 | Metrics: Classification | Model Development |
| Full Contingency Table against Reference (includes True/False Positives/Negatives) | 0.00% | 12 | 0 | 2 | Metrics: Classification | Model Development |
| True Positive (TP) | 8.33% | 12 | 1 | 1 | Metrics: Classification | Model Development |
| True Negative (TN) | 0.00% | 12 | 0 | 1 | Metrics: Classification | Model Development |
| False Negative / False Negative Rate | 8.33% | 12 | 1 | 2 | Metrics: Classification | Model Development |
| False Discovery Rate | 0.00% | 12 | 0 | 1 | Metrics: Classification | Model Development |
| False Omission Rate | 0.00% | 12 | 0 | 1 | Metrics: Classification | Model Development |
| F score / Dice Coefficient | 0.00% | 12 | 0 | 1 | Metrics: Classification | Model Development |
| NNT | 0.00% | 12 | 0 | 1 | Metrics: Utility | Utility Assessment |
| Net Benefit (Decision Curve) | 0.00% | 12 | 0 | 3 | Metrics: Utility | Utility Assessment |
| Relative Utility (Decision Curve) | 0.00% | 12 | 0 | 1 | Metrics: Utility | Utility Assessment |
| Net Reclassification Improvement | 0.00% | 12 | 0 | 5 | Metrics: Compare Two Model Discrimination | Model Development |
| Integrated Discrimination Improvement | 0.00% | 12 | 0 | 2 | Metrics: Compare Two Model Discrimination | Model Development |
| Compare model's performance to that of a baseline model, with statistical significance. | 0.00% | 12 | 0 | 2 | Comparison Against Baseline Model | Model Development |
| Specify what directions, explanations and other user-facing materials there will be with the model. | 0.00% | 12 | 0 | 9 | Intended Use: User | Use Case |
| Description of the clinical activity or annoyance that may be required to make the model, e.g. manually enter info, move to another screen, or otherwise make additional "clicks"? | 8.33% | 12 | 1 | 1 | Intended Use: Warnings | Use Case |
| A warning on when to stop use of model | 8.33% | 12 | 1 | 1 | Intended Use: Warnings | Use Case |
| Guidance on specific technical issues to address for integration of the model into your care setting, e.g. hardware, cloud, software or computing environment needs. | 8.33% | 12 | 1 | 2 | Deployment | Practical Feasibility |
| How private data from participants on which the model is deployed is protected. (this is deployment data, not training data) | 0.00% | 12 | 0 | 3 | Deployment: Tests: Data | Practical Feasibility |
| Check that no feature costs too much to have (e.g. dependencies, latency, instability, maintenance costs) compared with its added predictive value | 0.00% | 12 | 0 | 1 | Deployment: Tests: Data | Practical Feasibility |
| Programmatically enforce that input features adhere to meta-level requirements (e.g. deprecated features, protected features). | 0.00% | 12 | 0 | 1 | Deployment: Tests: Data | Deployed Model: Execution |
| In deployment, a new feature can be added quickly (e.g. within 1-2 months) to the model from ideation. | 0.00% | 12 | 0 | 1 | Deployment: Tests: Data | Deployed Model: Execution |
| Develop unit tests for input features. | 0.00% | 12 | 0 | 1 | Deployment: Tests: Data | Deployed Model: Monitoring |
| Monitor input data to ensure that it falls within correct ranges and invariances. | 0.00% | 12 | 0 | 1 | Deployment: Tests: Data | Deployed Model: Monitoring |
| Perform unit tests of model specification: API usage (e.g. check API calls on a random input) and algorithmic correctness (is it producing the predictions for the correct reasons)? | 0.00% | 12 | 0 | 1 | Deployment: Tests: Infrastructure | Deployed Model: Monitoring |
| Continuously perform an integration test: a fully automated test that runs regularly and uses the entire pipeline, validating that data and code can successfully move through each stage and that the resulting model performs well. | 0.00% | 12 | 0 | 1 | Deployment: Tests: Infrastructure | Deployed Model: Monitoring |
| Model allows debugging by step-by-step computation of training/inference on single example | 0.00% | 12 | 0 | 1 | Deployment: Tests: Infrastructure | Deployed Model: Monitoring |
| Models are tested via a canary process before they enter production serving environments: | 0.00% | 12 | 0 | 1 | Deployment: Tests: Infrastructure | Deployed Model: Monitoring |
| Every model specification undergoes a code review and is checked in to a repository: | 0.00% | 12 | 0 | 1 | Deployment: Updating | Deployed Model: Monitoring |
| Check that models can be quickly, easily rolled back in case of emergency. | 0.00% | 12 | 0 | 1 | Deployment: Updating | Deployed Model: Monitoring |
| Monitor regressions in prediction quality in newer data. | 8.33% | 12 | 1 | 3 | Deployment: Monitoring | Deployed Model: Monitoring |
| Monitor the age of the model and determine how old will affect the staleness of the model. | 0.00% | 12 | 0 | 1 | Deployment: Monitoring | Deployed Model: Monitoring |
| The deployment team has a line of communication with upstream, dependent data sources and is familiar with new data source changes. | 0.00% | 12 | 0 | 1 | Deployment: Monitoring | Deployed Model: Monitoring |
| Programmatically check whether data matches invariants in schema and alert when they diverge significantly, tuning a reasonable false positive/false negative point. | 0.00% | 12 | 0 | 1 | Deployment: Monitoring | Deployed Model: Monitoring |
| Monitor numerical stability of model, including NaNs and infinities in model components/weights or predictions. | 0.00% | 12 | 0 | 2 | Deployment: Monitoring | Deployed Model: Monitoring |
| Check that training and serving features compute the same values, either by direct comparison of features computed in both systems, or by comparing distributions. | 0.00% | 12 | 0 | 1 | Deployment: Monitoring | Deployed Model: Monitoring |
| Check degradations in the model computational performance. | 0.00% | 12 | 0 | 1 | Deployment: Monitoring | Deployed Model: Monitoring |
| Acknowledge any multiplicity of analyses or comparisons which may cause spurious signals. | 0.00% | 12 | 0 | 2 | Limitations | Model Development |

#### **eTable 5: Rarely Reported Atoms**. All atoms reported by 10% or less of applicable Model Briefs are listed. Reporting Rate indicates the % of the Model Briefs that provided the information requested in the atom. Task and Stage indicate the atoms’ related task and related stage of clinical predictive model development, respectively ^1^.

| **Atom Description** | **# Consensus** | **Task** | **Stage** | **Reporting Rate** | **# Model Reporting Guidelines requesting** |
| --- | --- | --- | --- | --- | --- |
| Who and how to contact with questions about the model | 0 | Overview | Other: Personnel | 100.0% | 2 |
| Summarize, discuss and interpret results | 0 | Overview | Other | 91.7% | 2 |
| Specify who funded / supported the study and clarify any conflicts of interest | 0 | Overview | Other: Personnel | 0.0% | 4 |
| It is clear if candidate predictors are available at time of intended use of model? | 0 | Data Composition: Input | Practical Feasibility | 91.7% | 2 |
| Report the distribution of severity/stage of disease in those with the target condition | 0 | Data Composition: Output | Model Development | 0.0% | 1 |
| Report the distribution of alternative diagnoses in those without the target condition. | 0 | Data Composition: Output | Model Development | 8.3% | 1 |
| Describe the design of the study that was used to collect the data. | 0 | Study Design/Population | Model Development | 83.3% | 5 |
| Describe how participants were enrolled or recruited into the data. | 0 | Data Collection & Methods | Model Development | 58.3% | 3 |
| Define the timeline of data collection. This could, for example, include participant recruitment time, time of predictor measurement, and outcome measurement/followup time. | 0 | Data Collection & Methods | Model Development | 100.0% | 9 |
| Details of treatments received by participants, if relevant. (NOT studying specific interventions for patients, just what treatments they may be receiving already) | 0 | Data Collection & Methods | Model Development | 90.9% | 2 |
| Overview of data collection, annotation, and quality process | 0 | Data Collection & Methods | Model Development | 66.7% | 8 |
| Describe the annotation process of the input data, including who annotated the input data, what instructions they were given, and what expertise was needed. | 0 | Data Collection & Methods: Input | Model Development | 18.2% | 4 |
| Blinding of Data Collectors/Predictor Assessors to outcomes, if done | 0 | Data Collection & Methods: Input | Model Development | 0.0% | 4 |
| Describe the annotation process of the output data, including who annotated the output data, what instructions they were given, and what expertise was needed. | 0 | Data Collection & Methods: Outcome | Model Development | 27.3% | 7 |
| Blinding of Outcome Assessors to predictors of the model, if done | 0 | Data Collection & Methods: Outcome | Model Development | 0.0% | 7 |
| Consistent Outcome Definition and Measurement for all patients | 0 | Data Collection & Methods: Outcome | Model Development | 100.0% | 3 |
| Reference standard for determining the outcome, if used | 0 | Data Collection & Methods: Outcome | Model Development | 44.4% | 3 |
| If there was a time interval and any interventions that occurred between the diagnostic index test and the reference standard, report it. | 0 | Data Collection & Methods: Outcome | Model Development | 57.1% | 1 |
| Predictors Not Part of Outcome (e.g. in panel or consensus diagnosis) | 0 | Data Collection & Methods: Outcome | Model Development | 100.0% | 2 |
| The time interval between the assessment of the predictors and the outcome is appropriate to allow the correct type and representative number of relevant outcomes to be recorded | 0 | Data Collection & Methods: Outcome | Model Development | 91.7% | 1 |
| How data was preprocessed (data cleaning, predictor transformation, outlier removal, predictor coding) | 0 | Preprocessing and Data Cleaning | Model Development | 100.0% | 10 |
| It is clear if categorical predictors have been dichotomized or categorized prior to model development. | 0 | Preprocessing and Data Cleaning | Model Development | 83.3% | 2 |
| Were all enrolled participants included in the analysis? (Not doing so leads to risk of bias. Number of participants included in each analysis and whether the analysis was by original assigned groups) | 0 | Preprocessing and Data Cleaning | Model Development | 58.3% | 3 |
| If feature selection involved computing univariate associations between input features and outcomes (not recommended), document this. | 0 | Preprocessing and Data Cleaning | Model Development | 18.2% | 4 |
| Describe which features were allowed interactions. | 0 | Model Building | Model Development | 0.0% | 2 |
| If a survival function is used, provide the baseline survival function. | 0 | Model Summary | Model Formulation | N/A | 2 |
| Report some examination of what the model is doing beyond the primary performance measure. | 0 | Model Examination | Model Development | 75.0% | 7 |
| Prognostic Index Plot for Validation Data Set | 0 | Metrics: Discrimination | Model Development | 0.0% | 1 |
| D-statistic | 0 | Metrics: Goodness-of-Fit | Model Development | 0.0% | 3 |
| For survival curves, the log-rank test | 0 | Metrics: Goodness-of-Fit | Model Development | N/A | 1 |
| Survival Curve/Kaplan-Meier Curve superimposition (for Cox models) | 0 | Metrics: Calibration | Model Development | N/A | 2 |
| Acknowledge if the model is intended to inform decisions about human life or safety. | 0 | Ethics | Use Case | 91.7% | 1 |
| Discuss any limitations and caveats of the study. | 0 | Limitations | Use Case | 83.3% | 6 |
| Discuss if or why well-known predictors were omitted from the model. | 0 | Limitations | Model Development | 25.0% | 2 |

#### **eTable 6: Low Consensus Atoms.** All atoms with no consensus among the reviewers are listed. Reporting Rate indicates the % of the Model Briefs that provided the information requested in the atom; N/A means the atom did not apply to any Model Briefs. Task and Stage indicate the atoms’ related task and related stage of clinical predictive model development, respectively ^1^.

|  | **EPIC MODEL BRIEFS** | | | | | | | | | | | |
| --- | --- | --- | --- | --- | --- | --- | --- | --- | --- | --- | --- | --- |
|  | **Deterioration Index** | **Early Detection of Sepsis** | **Risk of Unplanned Readmission** | **Risk of Patient No-Show** | **Pediatric Risk of Hospital Admission or ED Visit** | **Risk of Hospital Admission or ED Visit** | **Inpatient Risk of Falls** | **Projected Block Utilization** | **Remaining Length of Stay** | **Risk of Admission of Heart Failure** | **Risk of Hospital Admission or ED Visit for Asthma** | **Risk of Hypertension** |
| **# Reported** | 77 | 68 | 76 | 73 | 53 | 81 | 66 | 55 | 68 | 64 | 62 | 66 |
| **# Applicable** | 166 | 169 | 169 | 170 | 171 | 173 | 171 | 171 | 173 | 172 | 173 | 173 |
| **Completion Rate** | 46% | 40% | 45% | 43% | 31% | 47% | 39% | 32% | 39% | 37% | 36% | 38% |
| **# Reported, excluding performance metrics** | 72 | 61 | 71 | 69 | 48 | 76 | 64 | 49 | 63 | 60 | 58 | 62 |
| **# Applicable, excluding performance metrics** | 140 | 144 | 143 | 144 | 145 | 147 | 145 | 145 | 147 | 146 | 147 | 147 |
| **Completion Rate, excluding performance metrics** | 51% | 42% | 50% | 48% | 33% | 52% | 44% | 34% | 43% | 41% | 39% | 42% |

#### **eTable 7: Epic Model Brief Completion Rates.** A Model Brief’s “completion rate” of a given group of atoms is the number of atoms reported by the Model Brief divided by the number of atoms that were applicable to that Model Brief. Cells are colored green if above 50% and yellow if between 25% and 50%.
